# Spread of SARS-CoV-2 Delta variant infections bearing the S:E484Q and S:T95I mutations in July and August 2021 in France

**DOI:** 10.1101/2021.09.13.21263371

**Authors:** Laura Verdurme, Gonché Danesh, Sabine Trombert-Paolantoni, Mircea T. Sofonea, Valérie Noel, Vincent Foulongne, Brigitte Montès, Edouard Tuaillon, Stéphanie Haim-Boukobza, Bénédicte Roquebert, Samuel Alizon

## Abstract

Analysing 92,598 variant screening tests performed on SARS-CoV-2 positive samples collected in France between 1 July and 31 August 2021 shows an increase of Kappa-like infections. Full genome sequencing reveals that these correspond to Delta variants bearing the S:E484Q mutation. Most of these sequences belong to a phylogenetic cluster and also bear the S:T95I mutation. Further monitoring is needed to determine if this trend is driven by undocumented superspreading events or an early signal of future viral evolutionary dynamics.

---

The emergence of SARS-CoV-2 variants shows that virus evolution can threaten our ability to control epidemics [1,2,3] and lead to increased infection virulence [4]. Recently, the variant of concern Delta, which has a high replication rate and immune evasion properties [5], has caused epidemic rebounds in several countries including France, where it was shown to have a 52 to 110% transmission advantage over the Alpha variant [6]. Ongoing surveillance protocols involve variant-specific screening tests that target key mutations and full genome sequencing. In particular, the extension of full vaccination coverage with 2 doses, which now reaches on average 62% in France and 59% in the European Union [7], can be expected to modify the virus fitness landscape and favour the spread of escape mutations [8].

## Tests and study population

We analysed 92,598 reverse transcriptase polymerase chain reaction (RT-PCR) variant-specific screening tests performed on samples collected in France by the CERBA network of medical analysis laboratories in patients from 5 to 80 years old between 1 July and 31 August 2021 (Table 1). 12.1% of these tests were performed using the VirSNiP SARS-CoV-2 Spike, TIB MOLBIOL assay, Berlin; 50.1% using the Variant Detect (TM) SARS-CoV-2 RT-PCR, PerkinElmer assay, USA, and 37.8% using the ID (TM) SARS-CoV-2/VOC evolution Pentaplex, ID Solution, France. The three tests screen for the presence of the E484K, the E484Q, and the L452R mutations in the Spike protein. Infections caused by the Delta variant are expected to be associated with L452R alone. The metadata associated with the variant-specific RT-PCR data is shown in Table 1.

**Table 1:** Description of the population studied with the N= 92,598 variant-specific screening tests performed between 1 July and 31 August 2021. CI: confidence interval.

| Variable | Level | Value |
| --- | --- | --- |
| Assay | ID Solution | 34,977 (37.8%) |
|  | PerkinElmer | 46,429 (50.1%) |
|  | TIB MOLBIOL | 11,192 (12.1%) |
| Sampling date | *median (95% CI)* | Aug 6 (Jul 8-Aug 30) |
| Age | *median (95% CI)* | 31 (8-72) |
| Sample origin | Hospital | 2,670 (2.9%) |
|  | Non-Hospital | 89,928 (97.1%) |
| Test result | Alpha-like | 3,179 (3.4%) |
|  | Beta/Gamma/Eta-like | 364 (0.4%) |
|  | Delta-like | 67,560 (73.0%) |
|  | Kappa-like | 49 (0.1%) |
|  | other | 21446 (23.2%) |
| Region | Ile-de-France | 26,811 (30.3%) |
|  | Provence-Alpes-Côte d'Azur | 26,159 (29.6%) |
|  | Normandie | 6,937 (7.8%) |
|  | Corse | 6,594 (7.5%) |
|  | Nouvelle-Aquitaine | 5,754 (6.5%) |
|  | Occitanie | 4,935 (5.6%) |
|  | Hauts-de-France | 4,058 (4.6%) |
|  | other | 7,500 (8.1%) |

To improve the specificity of the analysis, all the screening tests bearing the mutation S:E484Q and with low-enough Ct values, as well as a subset of other tests, were sequenced using the CovidSeq(tm) amplicon-based NGS assay according to supplier recommendations (Illumina technology). The bioinformatic workflow was executed using Nanoware(tm) software (Life and Soft®). Briefly, the fastQ files generated by NGS were prefiltered by seqktk v1.3 to eliminate bad quality bases. Human reads were then discarded using bwa v.0.7.17. Reads assemblage (megahit v1.2.9) and alignment (blast v2.9.0) were performed. A variant calling (freebayes v1.3.1) and annotation (snpEff v5.0) was carried out and a consensus sequence was constructed (ivar v1.3.1). Overall, we analysed 364 original full genome sequences.

## Variant-screening test results

The majority of the screening tests (67,560, i.e. 73.0%) were consistent with an infection by a Delta variant (i.e. L452R+/E484Q-/E484K-). Fewer samples were consistent with the Alpha variant (3,179, i.e. 3.4%, L452R-/E484Q-/E484K-), the Beta/Gamma/Eta variants (364, i.e. 0.4%, L452R-/E484Q-/E484K+), and the Kappa variant (49, i.e. 0.1%, L452R+/E484Q+/E484K-). The number of tests analysed varied across French regions (Supplementary Figure 1).

To analyse the temporal trend associated with the frequency of a given variant, we used the methods described in [6] and performed a multinomial regression using the Delta-like variant as a reference (Table 2). The most striking effect we detected is that the few screening tests consistent with the Kappa variant appear to be strongly increasing with time in some French regions. The proportion of infections consistent with an Alpha variant was significantly decreasing over time in most French regions, which is consistent with the rapid spread of the Delta variant [6].

**Table 2:** Factors associated with the detection of potential SARS-CoV-2 variants, as assessed by relative risk ratios (RRR) using a multinomial log-linear model, France, 1 July-31 August 2021 (n=92,598). In the model, screening tests consistent with a Delta variant are the reference for the RRR. See [6] for details about the methods used. NS: non-signficant

|  |  | **Alpha** | **Beta/Gamma/Eta** | **Kappa** | **other** |
| --- | --- | --- | --- | --- | --- |
| age |  | NS (0.92-1) | NS (0.96-1.2) | NS (0.57-1.1) | NS (0.99-1) |
| sample origin | hospital (reference) |  |  |  |  |
|  | Non-hospital | NS (0.8-1.3) | NS (0.44-1.2) | NS (0.22-3.8) | 1.21 (1.1-1.3) |
| assay | ID Solution (reference) |  |  |  |  |
|  | PerkinElmer | 0.54 (0.5-0.6) | NS (0.7-1.4) | 0.23 (0.082-0.64) | 1.37 (1.3-1.4) |
|  | TIB MOLBIOL | 0.27 (0.23-0.32) | NS (0.86-1.9) | NS (0.32-3.8) | 1.91 (1.8-2) |
| interaction<br>between the<br>calendar date and<br>the administrative<br>region | Normandie | 0.26 (0.21-0.33) | 0.05 (0.037-0.081) | NS (0.25-10) | NS (0.87-1) |
|  | Auvergne-Rhône-Alpes | 0.28 (0.094-0.85) | 0.04 (0.011-0.12) | NS (2.2E-5-1.6E3) | NS (0.42-1) |
|  | Bourgogne-Franche-Comté | 0.1 (0.068-0.16) | 0.14 (0.036-0.56) | 6.67 (1.1-41) | NS (0.92-1.3) |
|  | Bretagne | 0.19 (0.14-0.26) | 0.08 (0.042-0.15) | 0.11 (0.013-0.86) | 1.41 (1.2-1.6) |
|  | Centre-Val de Loire | 0.6 (0.4-0.9) | 0.22 (0.07-0.68) | NS (0.01-24) | NS (0.8-1.1) |
|  | Corse | 0.33 (0.25-0.43) | 0.29 (0.12-0.69) | 5.45 (1.2-25) | 1.15 (1-1.3) |
|  | Grand Est | 0.15 (0.08-0.29) | 0.04 (0.015-0.13) | NS (0.00028-600) | NS (0.69-1.2) |
|  | Hauts-de-France | 0.17 (0.14-0.22) | 0.06 (0.037-0.083) | NS (0.047-10) | NS (0.89-1.1) |
|  | Ile-de-France | 0.17 (0.16-0.19) | 0.07 (0.052-0.084) | 6.89 (3.4-14) | 1.07 (1-1.1) |
|  | Nouvelle-Aquitaine | 0.16 (0.12-0.21) | 0.26 (0.091-0.72) | NS (0.48-18) | 0.78 (0.69-0.88) |
|  | Occitanie | 0.19 (0.14-0.25) | 0.05 (0.027-0.079) | NS (0.31-15) | 1.27 (1.1-1.4) |
|  | Provence-Alpes-Côte d'Azur | 0.41 (0.36-0.47) | 0.23 (0.15-0.36) | NS (0.22-3) | 1.18 (1.1-1.2) |
|  | other | 0.12 (0.031-0.43) | NS (0.0015-110) | NS (2.7E-8-2.9E5) | NS (0.35-1.1) |

## Doubling times and transmission advantage in the Parie area

Focusing on the tests consistent with a Kappa infection, the majority (33/49, i.e. 67%) originated from the Ile-de-France region (the Paris area) and the incidence globally increased from July to August 2021 (Supplementary Figure S2). Following earlier studies [9,10], we assessed the change in relative frequencies of Kappa-consistent infections compared to Delta-consistent infections in the Ile-de-France region during August 2021. The results show that the former had a transmission advantage over the latter of 45% (95% confidence interval: 31 – 60%) (Figure 1). Using an earlier starting date in July had little effect on the estimations (Figure S3).

**Figure 1:**
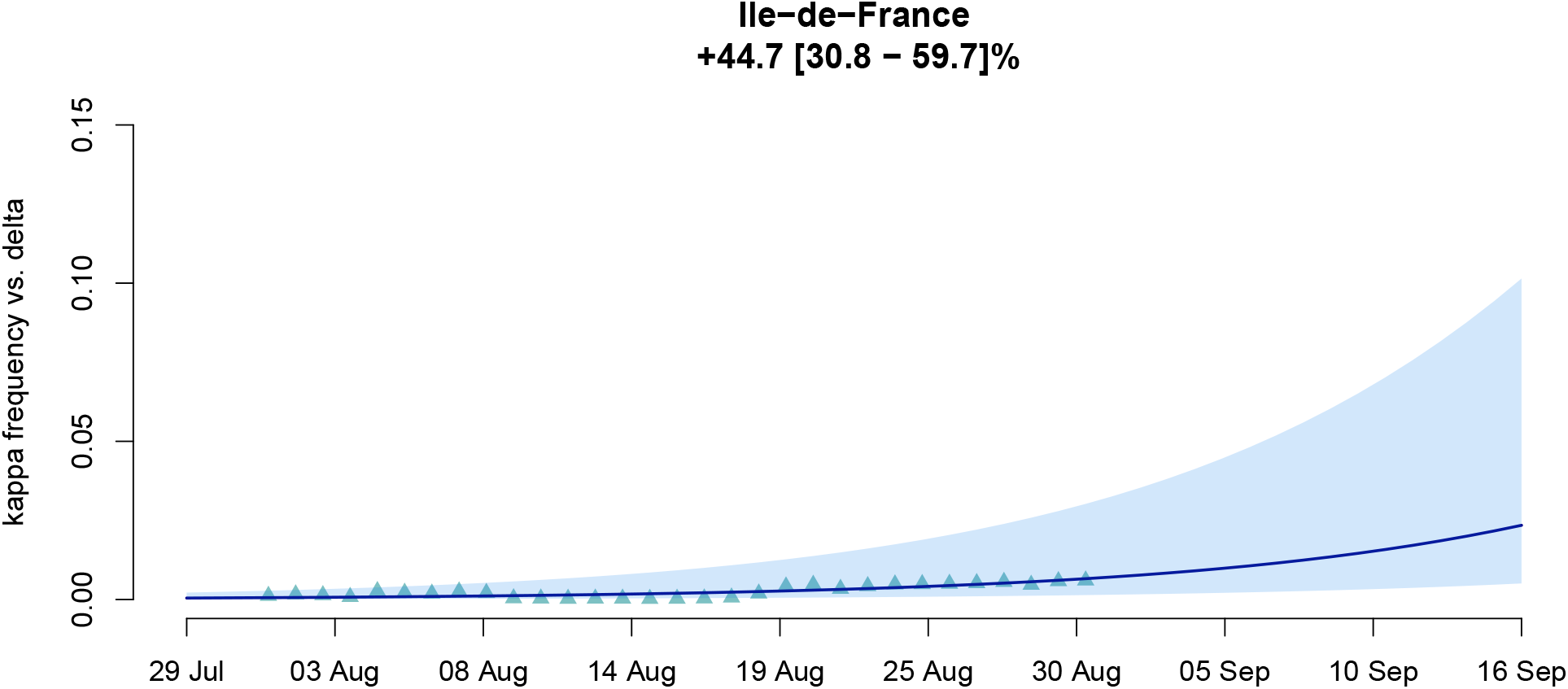
Estimated transmission advantage of the tests consistent with the Kappa variant with respect to that consistent with the Delta variant in Ile-de-France, 1-31 August 2021. The triangles indicate the generalised linear model (GLM)-fitted values and the line the output of the logistic growth model estimation. The shaded area represents the 95 %CI. The title indicates the estimated transmission advantage.

## Sequencing and phylogenetic analysis

Full genome sequencing and analysis through the nextclade pipeline [11] revealed that all the samples (35/35, 100%) bearing the S:E484Q mutation and identified as potential Kappa were in fact Delta variants bearing the E484Q mutation.

We then aligned the sequences using mafft [12], anonymised nucleotide position 23012 and performed a phylogenetic analysis using PhyML [13], using SMS to infer the substitution model [14]. The selected model was GTR+I+G. Strikingly, the majority of these Delta+E484Q sequences (28/35, 80%) clustered into the phylogeny into a clade (Figure 2). Most of these sequences from this clade (29/35, 83%) originated from individuals residing in the Ile-de-France region and the information was missing for 2 sequences. In addition to S:E484Q, the phylogenetic cluster is associated with the following substitutions that are not found in half of the other Delta sequences: ORF1a:T403I, ORF1a:D1167N, ORF1b:A1219S, and S:T95I. The latter is present in some variants of interest, such as Iota, and has been reported in vaccine breakthrough infections [15].

**Figure 2:**
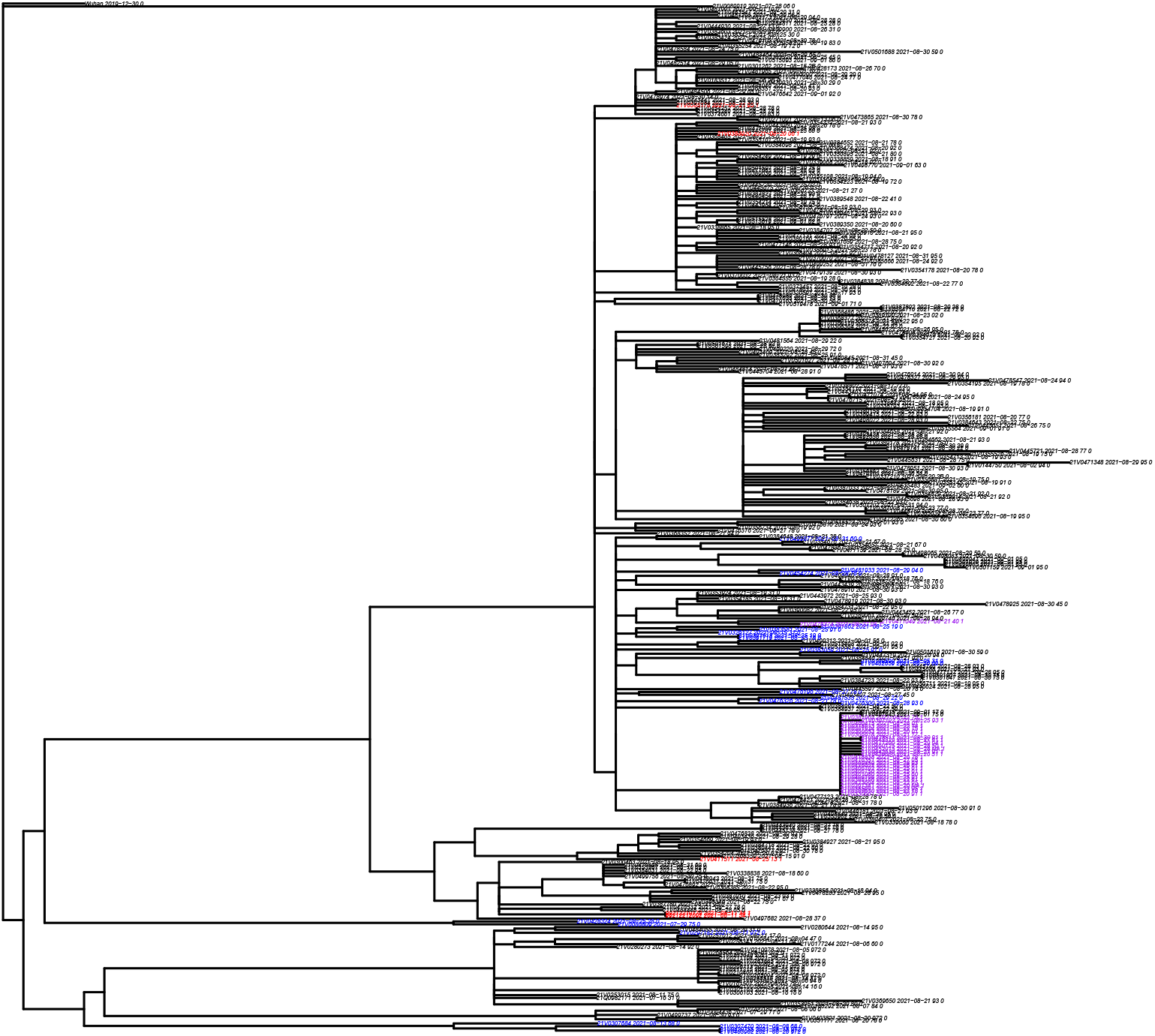
Phylogeny inferred from 364 Delta-positive sequences from France. The reference sequence from Wuhan (China) was used to root the phylogeny and align the sequences. Blue tips indicate sequences bearing the S:T95I mutations, red tips sequences with the S:E484Q mutation, and purple tips sequences with both mutations.

Finally, none of the individuals with Delta+E484Q reported travelling outside France 14 days before the sampling. As indicated above, nucleotide position 23,012, which is associated with the E484Q mutation, was ignored when performing the phylogenetic inference, meaning that the clustering originates from other mutations. Finally, based on the metadata analysis, there was no reported link between the 30 individuals from the phylogenetic cluster of interest (e.g. non were sequenced in the context of a transmission cluster). Note that two sequences in the cluster neither have the S:E484Q nor the S:T95I mutation. One possibility is the uncertainty in the phylogenetic reconstruction. Another could be the occurrence of genetic events such as recombination between two Delta lineages.

## Discussion

We report an increase in infections caused by Delta variants bearing the E484Q mutation in the Paris area. The significant transmission advantage we estimate for this mutation is consistent with it being associated with immune evasion [8] and with the high vaccination coverage in France [7]. Note that other lineages bearing a mutation in position S:484 were found to spread earlier in 2021 [16] but their transmission advantage was more limited (approximately 17% versus 35 to 63% here) and the reference strain was the Alpha variant, whereas is less contagious that the Delta variant [6] and has less immune evasion properties [5].

Even if we detect a significant increasing trend, the Delta+E484Q samples represent a very small proportion of the tests performed. Therefore, the transmission advantage could be driven by an undocumented superspreading event in the area of Paris in August, that would leave traces in virus genomes [17]. This could be consistent with the fact that the number of sequences from Delta+E484Q in the GISAID database is currently very low (935 of the 937,570 Delta sequences in GISAID [18]). Furthermore, combining these sequences with 8 Delta sequences per month (April to September) from Italy, Spain, France, UK, Belgium, and Germany did not identify a clear link with the phylogenetic cluster of interest (Supplementary Figure S4). *In vitro* studies using viral culture assays could be instrumental to understand this phenomenon. SARS-CoV-2 prevalence remains high and the beginning of the school term is likely to increase virus spread. Therefore, the alternative, more worrying, interpretation of these results, which is an adaptive advantage of a combination of the S:E484Q and S:T95I mutations, warrants close monitoring of these genomic patterns.

## Data Availability

Data will be made available after the peer-review process.

## Ethical statement

This study has been approved by the Institutional Review Board of the CHU of Montpellier and is registered at ClinicalTrials.gov with identifier NCT04738331.

## Acknowledgements

The authors thank Gilles Roullin, Eric Hedbaut and Virginie Dubois from CERBA for their technical assistance, and ETE team from CNRS, IRD, and University of Montpellier for discussion, and the EMERGEN consortium. This project was supported by the Occitanie region and the ANR (PHYEPI grant).

---

**Figure S1:**
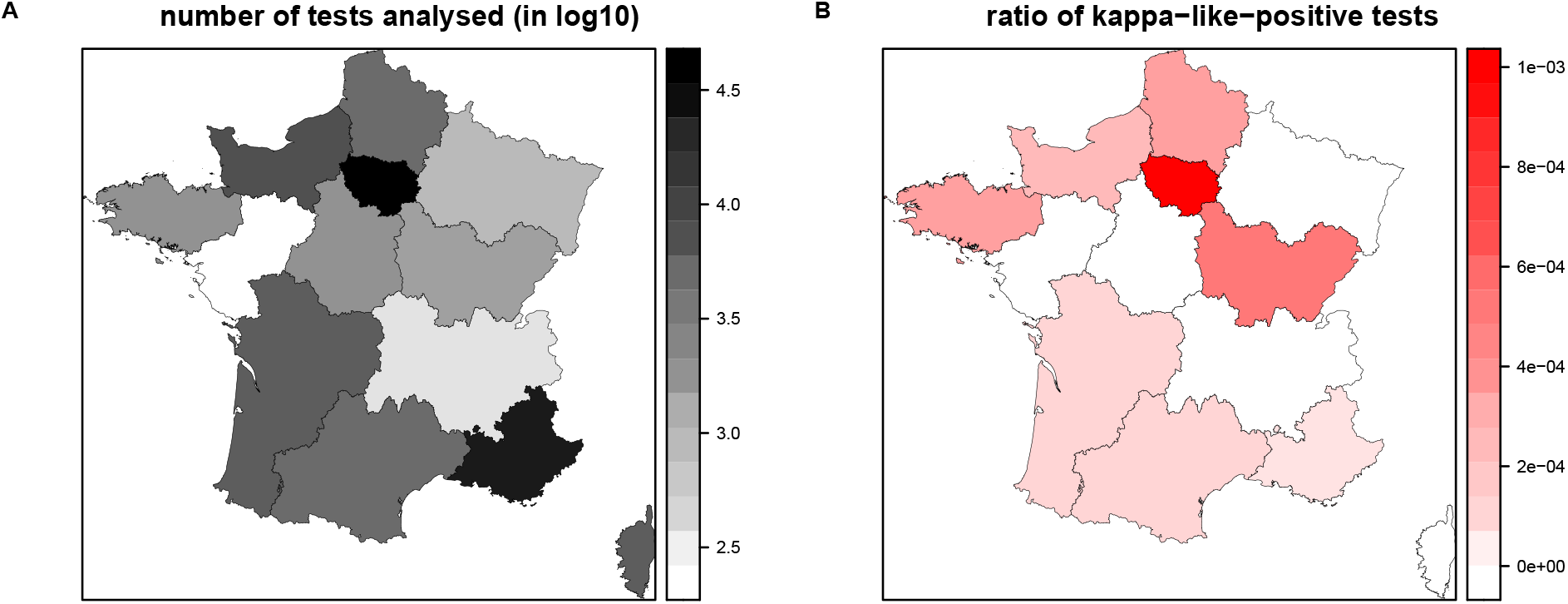
Number of tests analysed per French region between 1 July and 31 August 2021 (A) and ratio of these tests consistent with a Kappa infection (B).

**Figure S2:**
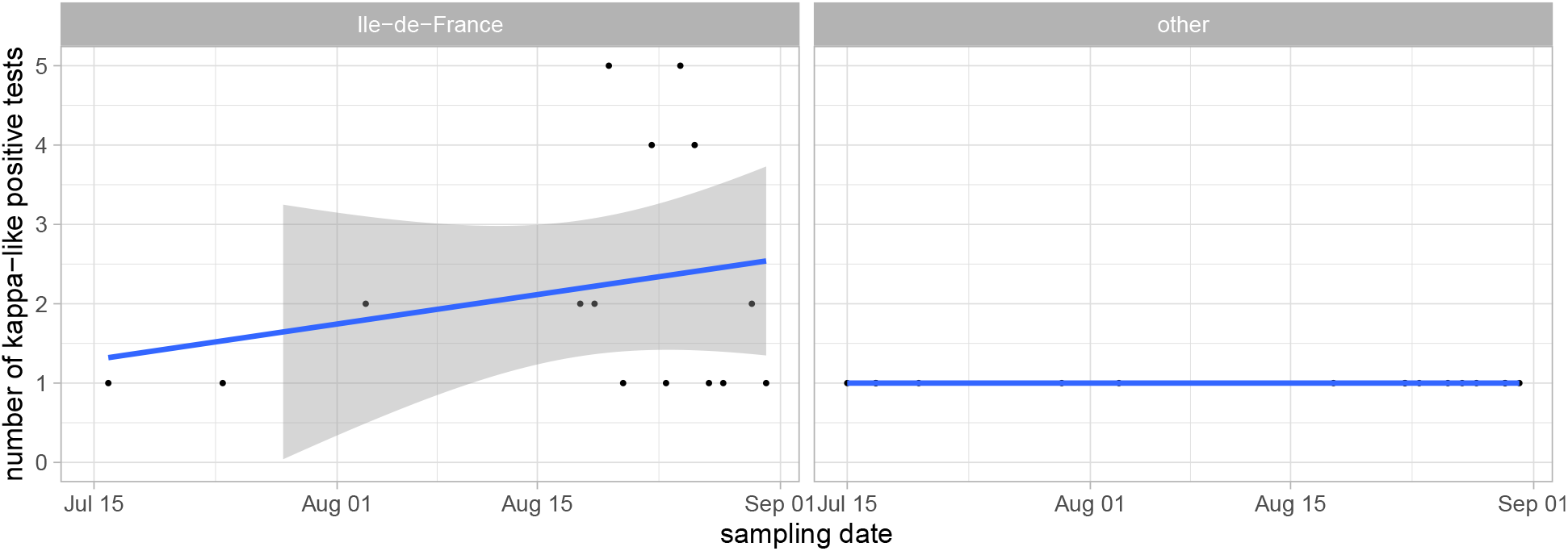
Daily number of tests consistent with a Kappa infection. Zeros are ignored. The lines show the output of a linear model and the shaded area the 95% confidence interval.

**Figure S3:**
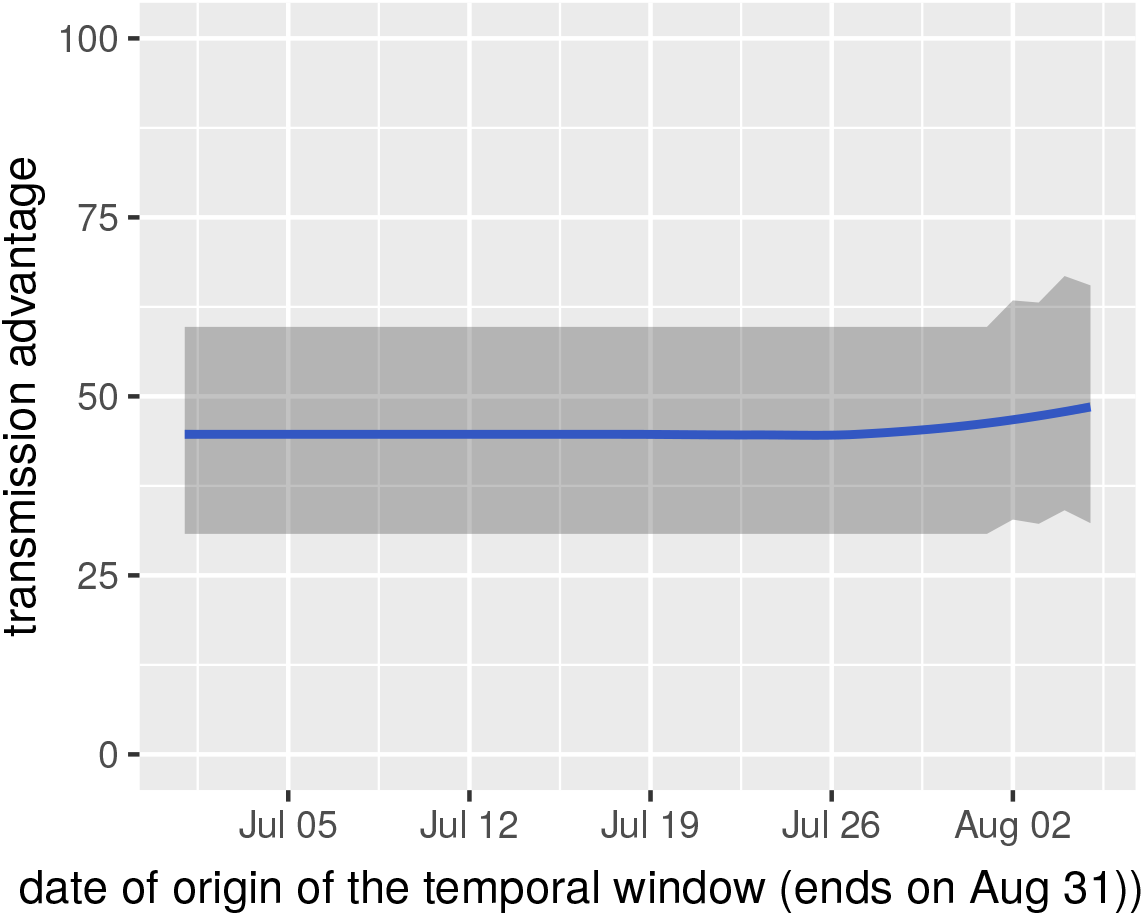
Effect of using an earlier date for the temporal interval on the estimates of the transmission advantage of the Kappa variant over the Delta variant. The starting date has little effect but the transmission advantage cannot be estimated on the whole time period (i.e. starting on July 1^st^), most likely because of stochastic variations during the month of July.

**Figure S4:**
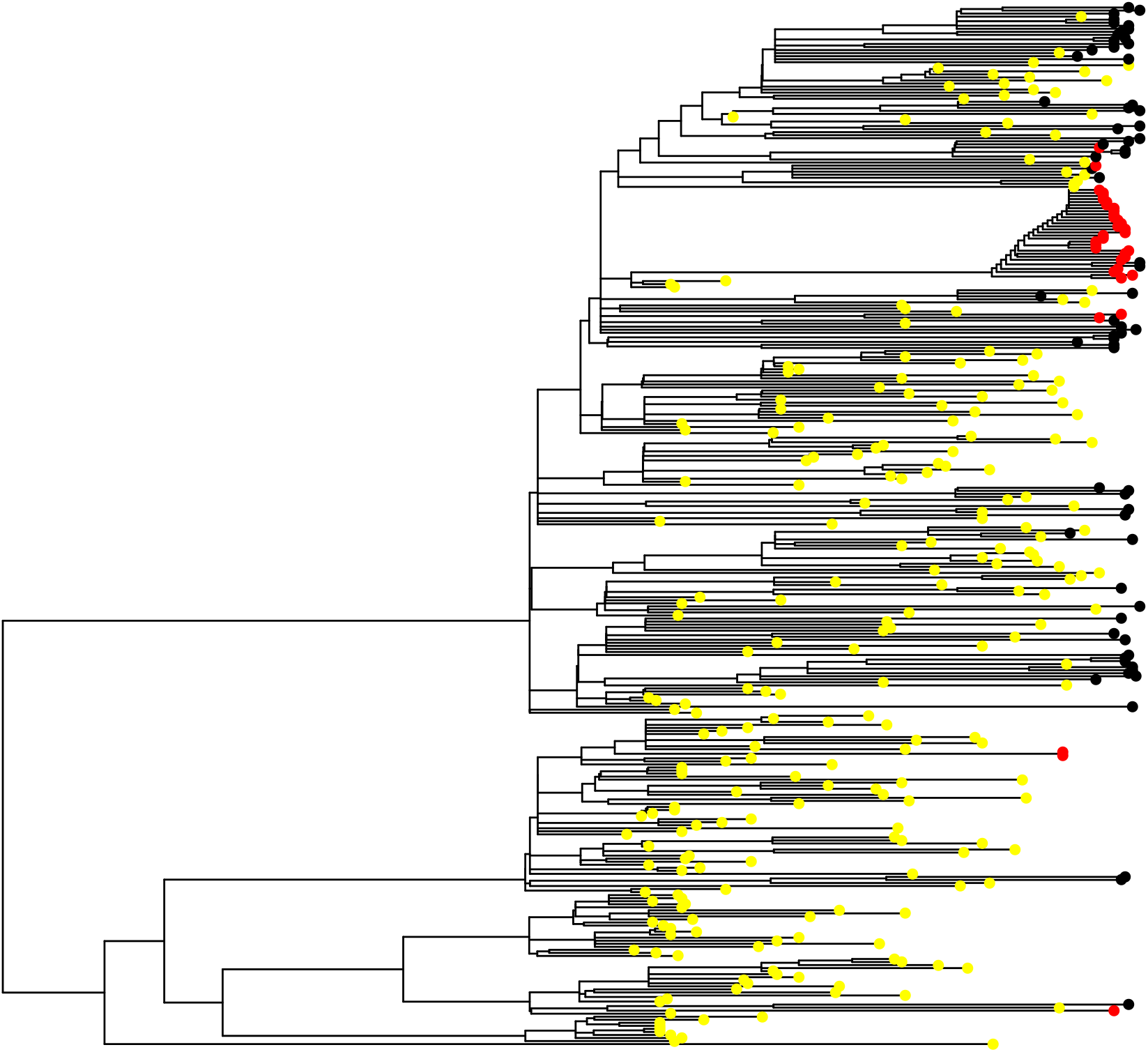
Phylogeny showing sequences from the European countries (in yellow) and the 364 original sequences from the study. Sequences from the study bearing the S:E484Q mutation are in red, the others are in black. GISAID accession numbers will be provided upon publications.

## References

[1] Davies NG, Abbott S, Barnard RC, Jarvis CI, Kucharski AJ, Munday JD, et al. Estimated transmissibility and impact of SARS-CoV-2 lineage B.1.1.7 in England. Science. 2021;eabg3055.

[2] Faria NR, Mellan TA, Whittaker C, Claro IM, Candido D da S, Mishra S, et al. Genomics and epidemiology of the P.1 SARS-CoV-2 lineage in Manaus, Brazil. Science. 2021;372(6544):815–21.

[3] Abdool Karim SS, de Oliveira T. New SARS-CoV-2 Variants — Clinical, Public Health, and Vaccine Implications. New England Journal of Medicine. 2021;384(19):1866–8.

[4] Alizon S, Sofonea MT. SARS-CoV-2 virulence evolution: avirulence theory, immunity, and trade-offs. Journal of Evolutionary Biology. 2021, in press, DOI 10.1111/jeb.13896

[5] Mlcochova P, Kemp S, Dhar MS, Papa G, Meng B, Ferreira IATM, et al. SARS-CoV-2 B.1.617.2 Delta variant replication and immune evasion. Nature. 2021; in press, DOI: 10.1038/s41586-021-03944-y.

[6] Alizon S, Haim-Boukobza S, Foulongne V, Verdurme L, Trombert-Paolantoni S, Lecorche E, et al. Rapid spread of the SARS-CoV-2 Delta variant in some French regions, June 2021. Eurosurveillance. 2021;26(28):2100573.

[7] Hannah Ritchie, Edouard Mathieu LR-G Cameron Appel, Charlie Giattino, Esteban Ortiz-Ospina, Joe Hasell, Bobbie Macdonald, Diana Beltekian, Roser M. Coronavirus pandemic (COVID-19). Our World in Data. 2020; https://ourworldindata.org/coronavirus Accessed September 9, 2021

[8] Starr TN, Greaney AJ, Addetia A, Hannon WW, Choudhary MC, Dingens AS, et al. Prospective mapping of viral mutations that escape antibodies used to treat COVID-19. Science. 2021; in press DOI: 10.1126/science.abf9302

[9] Chevin L-M. On measuring selection in experimental evolution. Biology Letters. 2011;7(2):210–3.

[10] Haim-Boukobza S, Roquebert B, Trombert-Paolantoni S, Lecorche E, Verdurme L, Foulongne V, et al. Detection of Rapid SARS-CoV-2 Variant Spread, France, January 26--February 16, 2021. Emerging Infectious Diseases. 2021;27(5):1496–9.

[11] Hadfield J, Megill C, Bell SM, Huddleston J, Potter B, Callender C, et al. Nextstrain: real-time tracking of pathogen evolution. Bioinformatics. 2018;34(23):4121–3.

[12] Katoh K, Rozewicki J, Yamada KD. MAFFT online service: multiple sequence alignment, interactive sequence choice and visualization. Briefings in Bioinformatics. 2019;20(4):1160–6.

[13] Guindon S, Gascuel O. A simple, fast, and accurate algorithm to estimate large phylogenies by maximum likelihood. Syst Biol. 2003;52(5):696–704.

[14] Lefort V, Longueville J-E, Gascuel O. SMS: smart model selection in PhyML. Molecular biology and evolution 2017: 34:2422–2424.

[15] Hacisuleyman E, Hale C, Saito Y, Blachere NE, Bergh M, Conlon EG, et al. Vaccine Breakthrough Infections with SARS-CoV-2 Variants. New England Journal of Medicine. 2021; in press DOI: 10.1056/NEJMoa2105000

[16] Roquebert B, Trombert-Paolantoni S, Haim-Boukobza S, Lecorche E, Verdurme L, Foulongne V, et al. The SARS-CoV-2 B.1.351 lineage (VOC β) is outgrowing the B.1.1.7 lineage (VOC α) in some French regions in April 2021. Eurosurveillance. 2021 Jun 10;26(23):2100447.

[17] Alizon S. Superspreading genomes. Science. 2021 Feb 5;371(6529):574–5.

[18] Latif A. A., Mullen J L., Alkuzweny M, Tsueng G, Cano M, Haag E, Zhou J, Zeller M, Hufbauer E, Matteson N, Wu C, Andersen K G, Su A I, Gangavarapu K, Hughes L D, and the Center for Viral Systems Biology. http://outbreak.info, https://outbreak.info/compare-lineages, Accessed 9 September 2021.

